# Is the Juice Worth the Squeeze? Overall Survival gain per unit treatment time as a metric of clinical benefit of systemic treatment in incurable cancers

**DOI:** 10.1101/2023.03.09.23287082

**Authors:** Nuradh Joseph, Vodathi Bamunuarachchi, Vimukthini Peiris, Sidath Wijeskera, Daminda Rajapakse, Sanjeeva Gunasekera

## Abstract

**Introduction:** Novel systemic therapeutic options such as enzyme inhibitors and monoclonal antibodies have transformed the practice of medical oncology in the recent past. However, survival gains remain modest in most cases. Quantifying the magnitude of benefit against financial and non-financial toxicity of treatment is pivotal in deciding treatment. We describe a novel metric which can be used to assess effectiveness novel therapeutics for incurable cancers.

**Methods:** The median overall survival was divided by the median duration of treatment to obtain the overall survival gain per treatment time which was the primary end-point of the study. This parameter was compared with the European Society of Medical Oncology Magnitude of clinical benefit scale (ESMO-MCBS) score. Spearman’s rank correlation coefficient was used to test the association between the novel metric and the ESMO-MCBS scores.

**Results:** Data were available for 30 drugs across 60 indications. The median overall survival per unit treatment time ranged from 0.68 (range 0.2-0.51). Only 18/60 indications had a ratio greater than 1 while 13/60 indications had a ratio less than 0.5. The median treatment duration was not mentioned in 11 indications and median progression free survival was substituted for the analysis. The ESMO-MCBS score was available for 49 of the indications. The Spearman’s rank correlation coefficient was 0.44575 and showed a statistically significant association between survival gain per unit treatment time and the ESMO-MCBS score (p = 0.00133).

**Conclusions:** Along with other metrics, the ratio of survival gain over treatment duration is a useful parameter to assess effectiveness of novel therapeutics in the palliative setting.

## Introduction

Although cure rates have improved significantly over the last couple of decades, nearly 50% of all cancers are still incurable (1). For patients with incurable cancer, the objective of treatment is to prolong survival while improving or maintaining quality of life. A large number of novel cancer therapeutic agents ranging from complex monoclonal antibodies to small molecule enzyme inhibitors and new types of hormonal treatment, have been approved for use in incurable cancers and while this has led to modest improvements in survival it comes at the price of significant toxicity as well as a heavy financial burden on health systems (2).

Oncologists prescribe systemic treatment in incurable cancer with a view to shrinking the tumour and/or preventing its growth but eventually the cancer develops resistance and progresses through treatment. The outcome end-points response rate, duration of response and progression free survival capture these aspects of treatment. However, they are not a robust surrogate marker for the overall survival which is considered as the primary end-point of choice (3,4).

Systemic treatment of incurable cancers is continued until tumour progression, which is defined as an increase in the sum of maximum tumour diameters of at least 20%, the development of any new lesions, or an unequivocal increase in non-measurable malignant disease, in comparison to the preceding assessment. Intolerable toxicity is another factor which could lead to a premature termination of treatment (5). The duration of treatment is an important metric that ought to be reported in all publications of trials involving novel cancer agents (6). It is the primary determinant of the direct cost of treatment and the time of exposure to treatment will impact on treatment toxicities.

In this work, we explore the novel metric overall survival gain per unit treatment time as a useful parameter to supplement other established measures of clinical benefit such the European Society of Medical Oncology - Magnitude of Clinical Benefit Scale (ESMO-MCBS) and the American Society of Clinical Oncology (ASCO) Value Framework score (7,8).

## Methods

Data on median overall survival gain and median duration of treatment were obtained from publications of phase III randomised clinical trials for novel anti-cancer therapeutics in the palliative setting.

In trials with significant treatment cross-over estimates of median survival gain were substituted where such data was available. Studies were excluded if no overall survival gain was demonstrated and where there were no published studies of estimates of overall survival gain. Trials in which median overall survival was not reached were also excluded from the analysis. If the median duration of treatment was not reported the median time to progression was substituted for it.

The median overall survival was divided by the median duration of treatment to obtain the overall survival gain per treatment time which was the primary end-point of the study. This parameter was compared with the ESMO-MCBS score. Spearman’s rank correlation coefficient was used to test the association between the novel metric and the ESMO-MCBS scores. The statistical Software R version 4.1.1 was used for analysis

## Results

Data were available for 30 drugs across 60 indications. Supplementary table S1 gives the full dataset of the analysis including the referenced publications from which data was extracted. Table 1 presents the values of the metric for 40 selected indications. The median overall survival was 5.8 months (range 1.4 – 42.2 months) and the median treatment duration was 8 months (rage, 1.9–78 months). The median overall survival per unit treatment time ranged from 0.68 (range 0.2–0.51). Supplementary Figure S1 depicts in the form of a histogram the number of indications for each survival gain per unit treatment time. Since the distribution was skewed a logarithmic transformation was performed to achieve greater normality, and the histogram of the logarithm of the metric is presented in Figure 1. The median of the logarithm of the survival gain per unit treatment time was −0.17 (range −0.71–0.7).

**Table 1.**
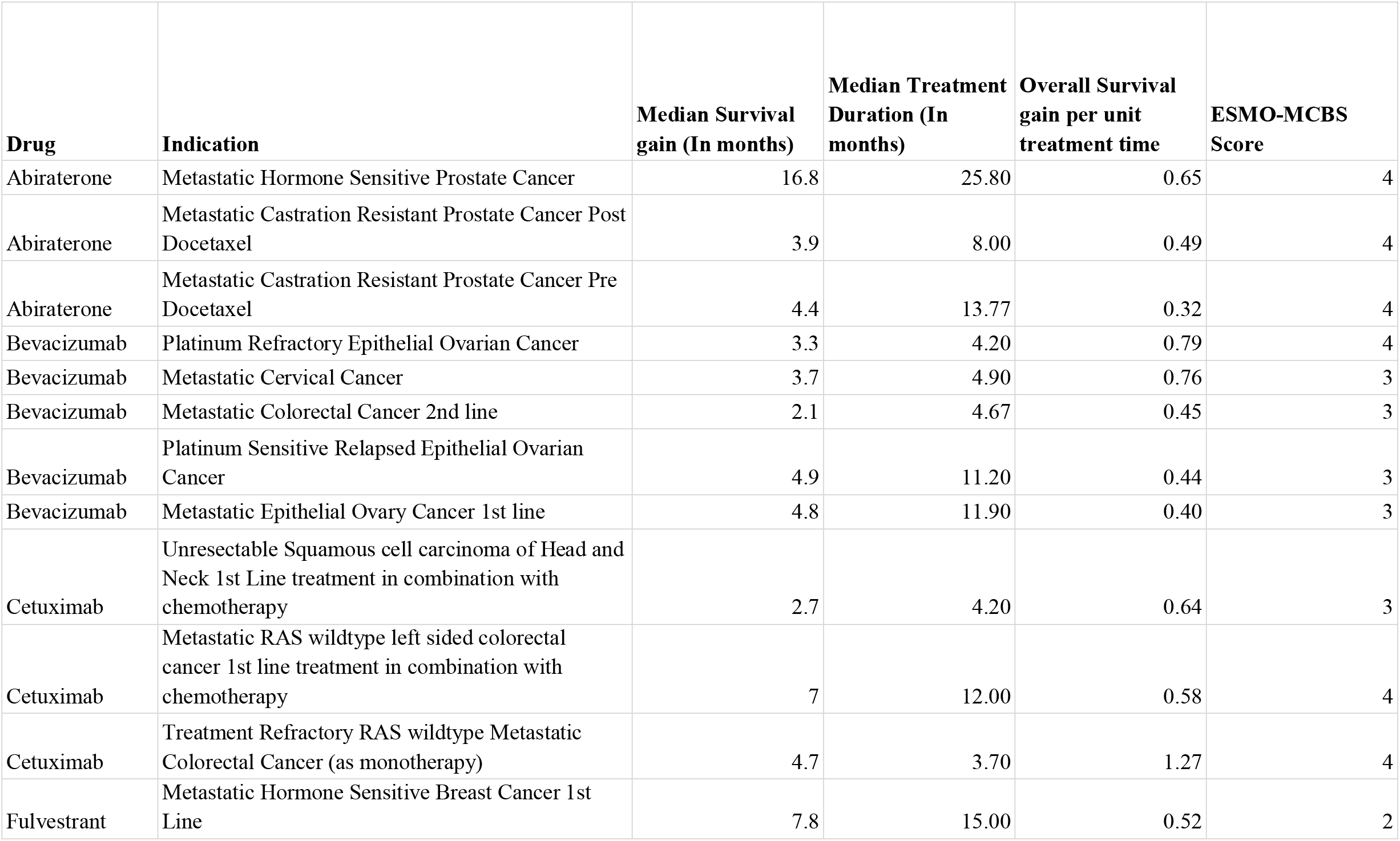
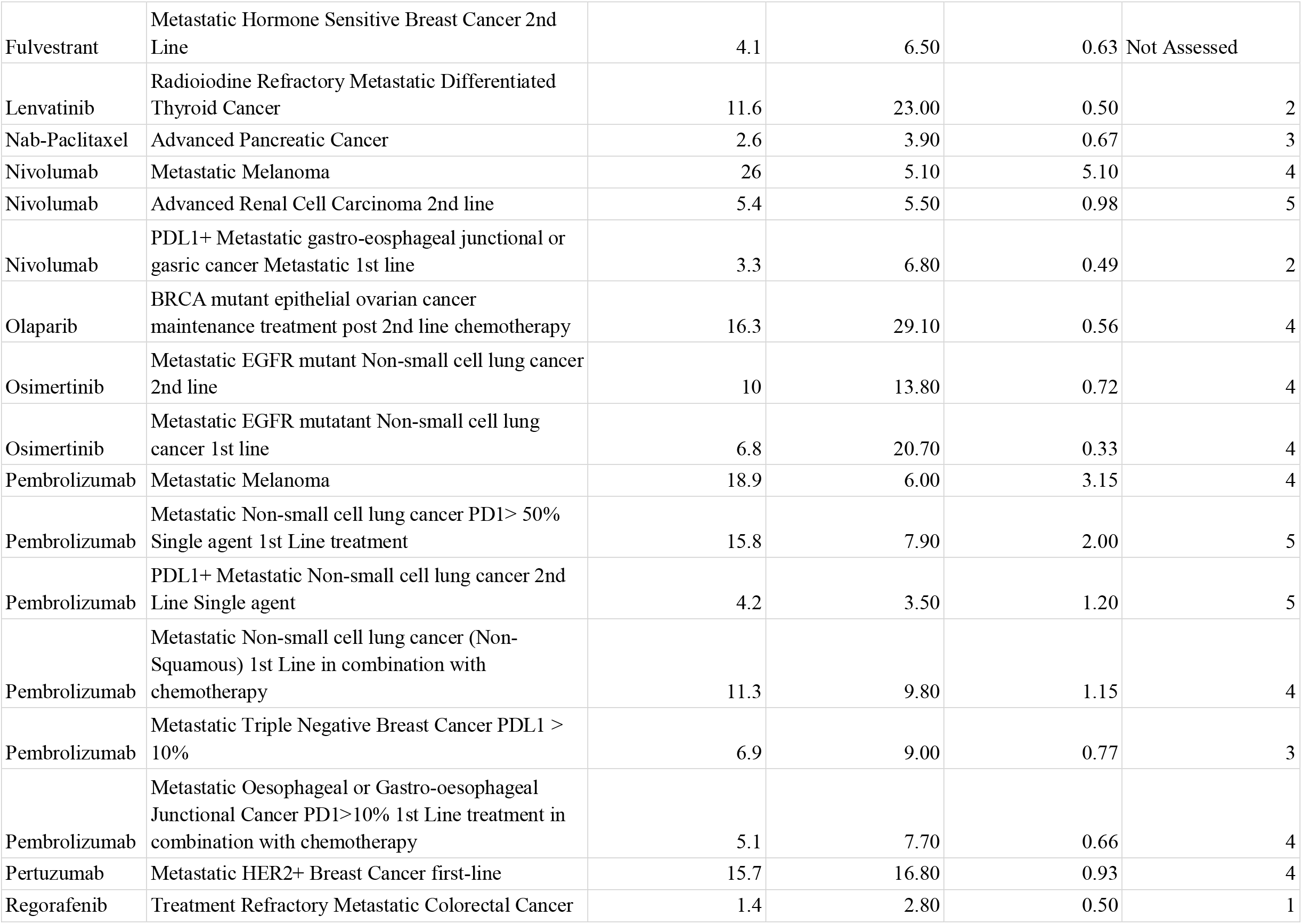
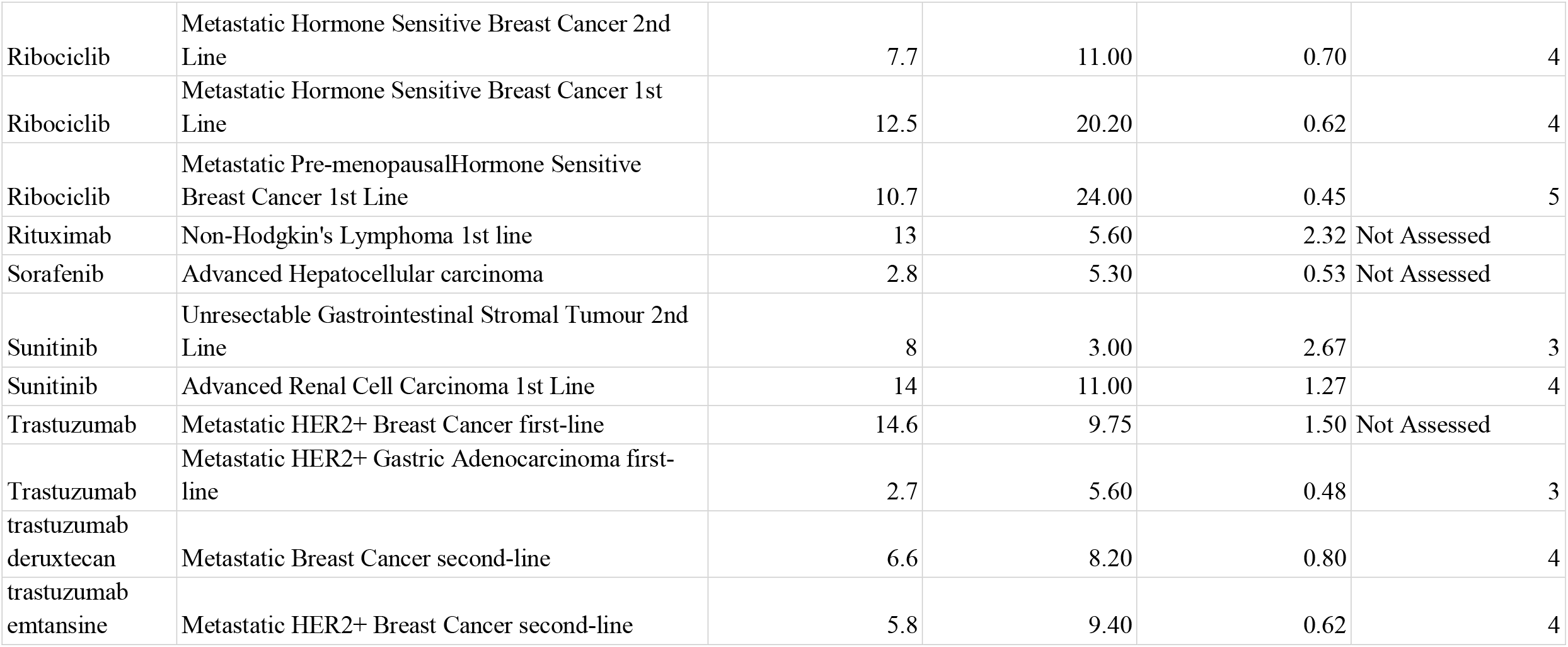
Overall survival gain per unit treatment time (selected indications)

| **Drug** | **Indication** | **Median Survival gain (In months)** | **Median Treatment Duration (In months)** | **Overall Survival gain per unit treatment time** | **ESMO-MCBS Score** |
| --- | --- | --- | --- | --- | --- |
| Abiraterone | Metastatic Hormone Sensitive Prostate Cancer | 16.8 | 25.80 | 0.65 | 4 |
| Abiraterone | Metastatic Castration Resistant Prostate Cancer Post Docetaxel | 3.9 | 8.00 | 0.49 | 4 |
| Abiraterone | Metastatic Castration Resistant Prostate Cancer Pre Docetaxel | 4.4 | 13.77 | 0.32 | 4 |
| Bevacizumab | Platinum Refractory Epithelial Ovarian Cancer | 3.3 | 4.20 | 0.79 | 4 |
| Bevacizumab | Metastatic Cervical Cancer | 3.7 | 4.90 | 0.76 | 3 |
| Bevacizumab | Metastatic Colorectal Cancer 2nd line | 2.1 | 4.67 | 0.45 | 3 |
| Bevacizumab | Platinum Sensitive Relapsed Epithelial Ovarian Cancer | 4.9 | 11.20 | 0.44 | 3 |
| Bevacizumab | Metastatic Epithelial Ovary Cancer 1st line | 4.8 | 11.90 | 0.40 | 3 |
| Cetuximab | Unresectable Squamous cell carcinoma of Head and Neck 1st Line treatment in combination with chemotherapy | 2.7 | 4.20 | 0.64 | 3 |
| Cetuximab | Metastatic RAS wildtype left sided colorectal cancer 1st line treatment in combination with chemotherapy | 7 | 12.00 | 0.58 | 4 |
| Cetuximab | Treatment Refractory RAS wildtype Metastatic Colorectal Cancer (as monotherapy) | 4.7 | 3.70 | 1.27 | 4 |
| Fulvestrant | Metastatic Hormone Sensitive Breast Cancer 1st Line | 7.8 | 15.00 | 0.52 | 2 |
| Fulvestrant | Metastatic Hormone Sensitive Breast Cancer 2nd Line | 4.1 | 6.50 | 0.63 | Not Assessed |
| Lenvatinib | Radioiodine Refractory Metastatic Differentiated Thyroid Cancer | 11.6 | 23.00 | 0.50 | 2 |
| Nab-Paclitaxel | Advanced Pancreatic Cancer | 2.6 | 3.90 | 0.67 | 3 |
| Nivolumab | Metastatic Melanoma | 26 | 5.10 | 5.10 | 4 |
| Nivolumab | Advanced Renal Cell Carcinoma 2nd line | 5.4 | 5.50 | 0.98 | 5 |
| Nivolumab | PDL1+ Metastatic gastro-esophageal junctional or gastric cancer Metastatic 1st line | 3.3 | 6.80 | 0.49 | 2 |
| Olaparib | BRCA mutant epithelial ovarian cancer maintenance treatment post 2nd line chemotherapy | 16.3 | 29.10 | 0.56 | 4 |
| Osimertinib | Metastatic EGFR mutant Non-small cell lung cancer 2nd line | 10 | 13.80 | 0.72 | 4 |
| Osimertinib | Metastatic EGFR mutant Non-small cell lung cancer 1st line | 6.8 | 20.70 | 0.33 | 4 |
| Pembrolizumab | Metastatic Melanoma | 18.9 | 6.00 | 3.15 | 4 |
| Pembrolizumab | Metastatic Non-small cell lung cancer PD1> 50% Single agent 1st Line treatment | 15.8 | 7.90 | 2.00 | 5 |
| Pembrolizumab | PDL1+ Metastatic Non-small cell lung cancer 2nd Line Single agent | 4.2 | 3.50 | 1.20 | 5 |
| Pembrolizumab | Metastatic Non-small cell lung cancer (Non-Squamous) 1st Line in combination with chemotherapy | 11.3 | 9.80 | 1.15 | 4 |
| Pembrolizumab | Metastatic Triple Negative Breast Cancer PDL1 > 10% | 6.9 | 9.00 | 0.77 | 3 |
| Pembrolizumab | Metastatic Oesophageal or Gastro-oesophageal Junctional Cancer PD1>10% 1st Line treatment in combination with chemotherapy | 5.1 | 7.70 | 0.66 | 4 |
| Pertuzumab | Metastatic HER2+ Breast Cancer first-line | 15.7 | 16.80 | 0.93 | 4 |
| Regorafenib | Treatment Refractory Metastatic Colorectal Cancer | 1.4 | 2.80 | 0.50 | 1 |
| Ribociclib | Metastatic Hormone Sensitive Breast Cancer 2nd Line | 7.7 | 11.00 | 0.70 | 4 |
| Ribociclib | Metastatic Hormone Sensitive Breast Cancer 1st Line | 12.5 | 20.20 | 0.62 | 4 |
| Ribociclib | Metastatic Pre-menopausalHormone Sensitive Breast Cancer 1st Line | 10.7 | 24.00 | 0.45 | 5 |
| Rituximab | Non-Hodgkin's Lymphoma 1st line | 13 | 5.60 | 2.32 | Not Assessed |
| Sorafenib | Advanced Hepatocellular carcinoma | 2.8 | 5.30 | 0.53 | Not Assessed |
| Sunitinib | Unresectable Gastrointestinal Stromal Tumour 2nd Line | 8 | 3.00 | 2.67 | 3 |
| Sunitinib | Advanced Renal Cell Carcinoma 1st Line | 14 | 11.00 | 1.27 | 4 |
| Trastuzumab | Metastatic HER2+ Breast Cancer first-line | 14.6 | 9.75 | 1.50 | Not Assessed |
| Trastuzumab | Metastatic HER2+ Gastric Adenocarcinoma first-line | 2.7 | 5.60 | 0.48 | 3 |
| trastuzumab<br>deruxtecan | Metastatic Breast Cancer second-line | 6.6 | 8.20 | 0.80 | 4 |
| trastuzumab<br>emtansine | Metastatic HER2+ Breast Cancer second-line | 5.8 | 9.40 | 0.62 | 4 |

**Figure 1.**
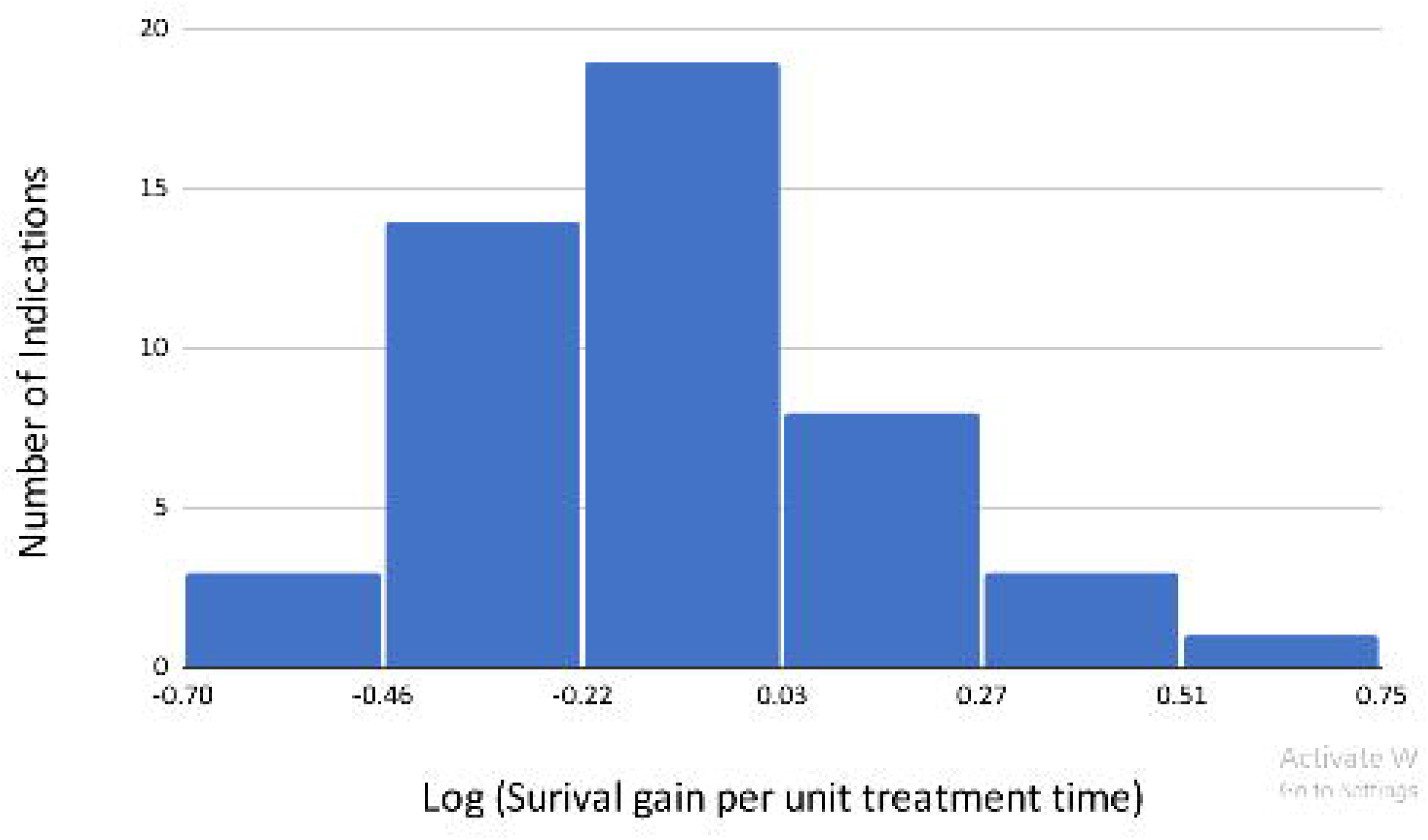
Distribution of indications against logarithm of survival gain per unit treatment time.

Only 18/60 indications had a ratio greater than 1 while 13/60 indications had a ratio less than 0.5. The median treatment duration was not mentioned in 11 indications and median progression free survival was substituted for the analysis. The ESMO-MCBS score was available for 49 of the indications. The Spearman’s rank correlation coefficient was 0.44575 and showed a statistically significant association between survival gain per unit treatment time and the ESMO-MCBS score (p = 0.00133).

## Discussion

In this work, we present a novel metric, survival gain per unit treatment time, as a useful parameter that could complement other measures of clinical benefit such as the ESMO-MCBS score. Ideally clinicians would want the benefit of treatment to outlast its duration, leaving a prolonged therapeutic legacy of benefit to the patient. This can be achieved if the novel drugs result in significant tumour shrinkage which is sustained for a prolonged time period. Treatments with a survival gain per treatment time > 1 would fall into this category, and are likely to be preferred by patients since the gain in survival is greater than exposure to the drug and its toxicity. In addition, they are likely to be more cost-effective since the cost of treatment is related to drug exposure time. A value of less than 0.5 with our novel metric, would indicate that patients need to be exposed to the treatment for almost twice the duration of survival gain, and these are likely to pose a heavy burden in terms of toxicity, both financial and non-financial for modest benefit.

A significant proportion of patients are not fit for second line treatment at the time of disease progression and there is a trend to use the most efficacious agents earlier in the course of the disease trajectory of metastatic cancer with a view to improving survival. The rationale for this approach is firmly rooted in the proportional hazards model, where for the same relative reduction of risk of death a higher absolute gain can be achieved if treatment is initiated early.

It was interesting to note that when considering first-line versus later line of treatment in the same disease for the same agent, it could be discerned that the survival gain per treatment time is higher when used in later lines of treatments for most drugs. Since treatment duration is often longer when used in the first line setting, it follows that the cost of treatment would also be greater in comparison to use of the same agent in the second line. The lower survival gain per treatment time in first line use would suggest that more robust biomarkers are needed to distinguish patients who would benefit from first line treatment from those who are suitable for sequential treatment. A notable exception to this trend was observed with the use of abiraterone in hormone sensitive metastatic prostate cancer where the survival gain per treatment time was greater in this setting than in the castration resistant phases of the disease.

Our metric places a higher premium on indications for which there are existing treatment options, since the novel agent has to achieve a survival gain relative to its treatment duration. We believe that this is a strength of the metric since it helps select treatments which are robustly effective from those which achieve only modest gains.

Since its publication the ESMO-MCBS has found widespread application as a robust tool of stratifying the benefit of each novel agent since it weaves together improvements in prolongation of survival while considering toxicity and quality of life into a single parameter. In the palliative setting the ESMO-MCBS classifies reported outcomes into five levels 1-5 with higher values indicating a higher clinical benefit. For trials reporting improvements in overall survival, the ESMO-MBCS scoring is based upon a consideration of the absolute gain in median overall survival, the lower limit of the confidence interval of the hazard ratio as well as reported gains in quality of life as well reduced grade 3 or 4 toxicity in comparison to the control arm.

It has been shown that although the cost of novel cancer drugs has increased over the past decade, clinical benefits did not follow a similar trend (9). Although there was a statistically significant association between our metric and the ESMO-MBCS score. The highest values for the novel metric was seen with the use of the PD-1 inhibitors nivolumab and pembrolizumab in metastatic melanoma, for which the ESMO-MBCS score was 4. However, while ribociclib in premenopausal metastatic breast cancer had a maximum value of 5 within the ESMO-MBCS scoring, it had a low value of 0.45 with our metric. Sunitinib in the second line treatment of unresectable gastrointestinal stromal tumours had a high value of 2.67 in our study but had an ESMO-MBCS score of just 3.

Unlike the ESMO-MBCS which is reported as a score of an ordinal scale, our metric is a continuous variable. Reporting the survival gain per treatment duration along with the conventional end-points such as response rates, median overall survival and the ESMO-MBCS would enable clinicians and patients to make more informed therapeutic decisions especially when weighing the benefit against toxicity.

It needs to be emphasised that the median duration of treatment was not disclosed in 11/60 studies that were screened. As mentioned previously, treatment duration is pivotal in deciding the cost of treatment which is an integral component of all cost-effectiveness and health technology modelling studies, and its reporting needs to be made mandatory in publications of phase III randomised trials.

There are a number of limitations in our study and in our novel metric. First, we excluded studies in which no overall survival gain was established. In many trials, when the primary end-point of a gain in progression-free survival is met crossover is permitted, adding to the complexity of determining the impact of the novel drug on overall survival (10,11). It would also mean that treatments that achieve improvements in quality of life without prolonging survival were also not considered in this analysis (12). Second, we used median progression-free survival as a surrogate in studies which did not report the median duration of treatment. As mentioned before, treatment in the non-curative setting is continued until disease progression or development of treatment toxicity and as such, progression free survival would therefore be longer than the duration of treatment in most instances. This may have led to underestimation of the survival gain per unit treatment time for certain indications.

## Conclusion

Overall survival gain per unit treatment duration is a potentially useful metric that would provide vital insights in quantifying the benefit of treatment of novel cancer therapeutics in the palliative setting. The juice in the form of survival gain must certainly do justice to the squeeze both in terms of financial and somatic toxicities.

## Data Availability

All data produced in the present work are contained in the manuscript

## Data Availability

All data produced in the present work are contained in the manuscript

## List of Tables and Figures

**Supplementary Table 1.**
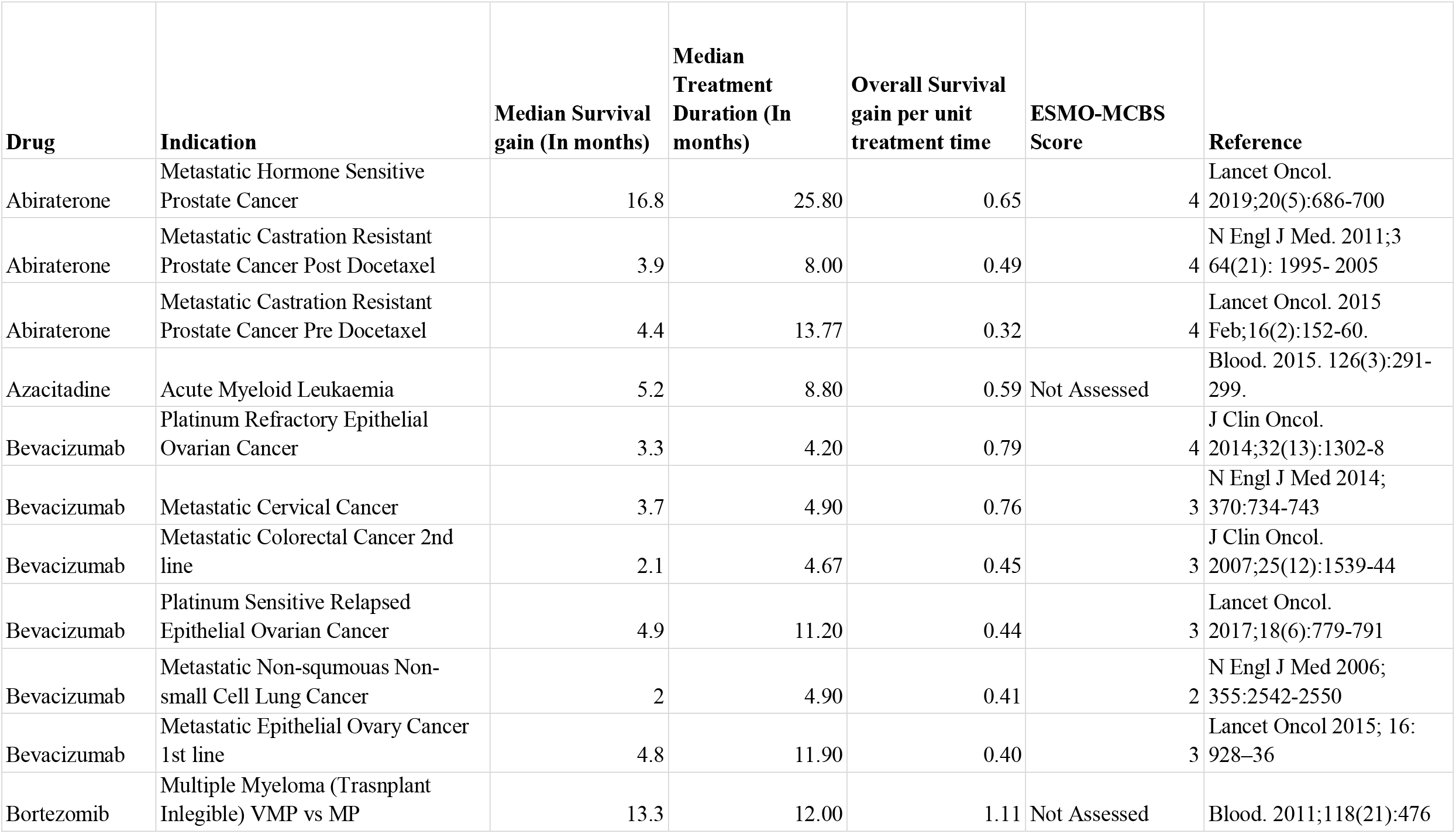
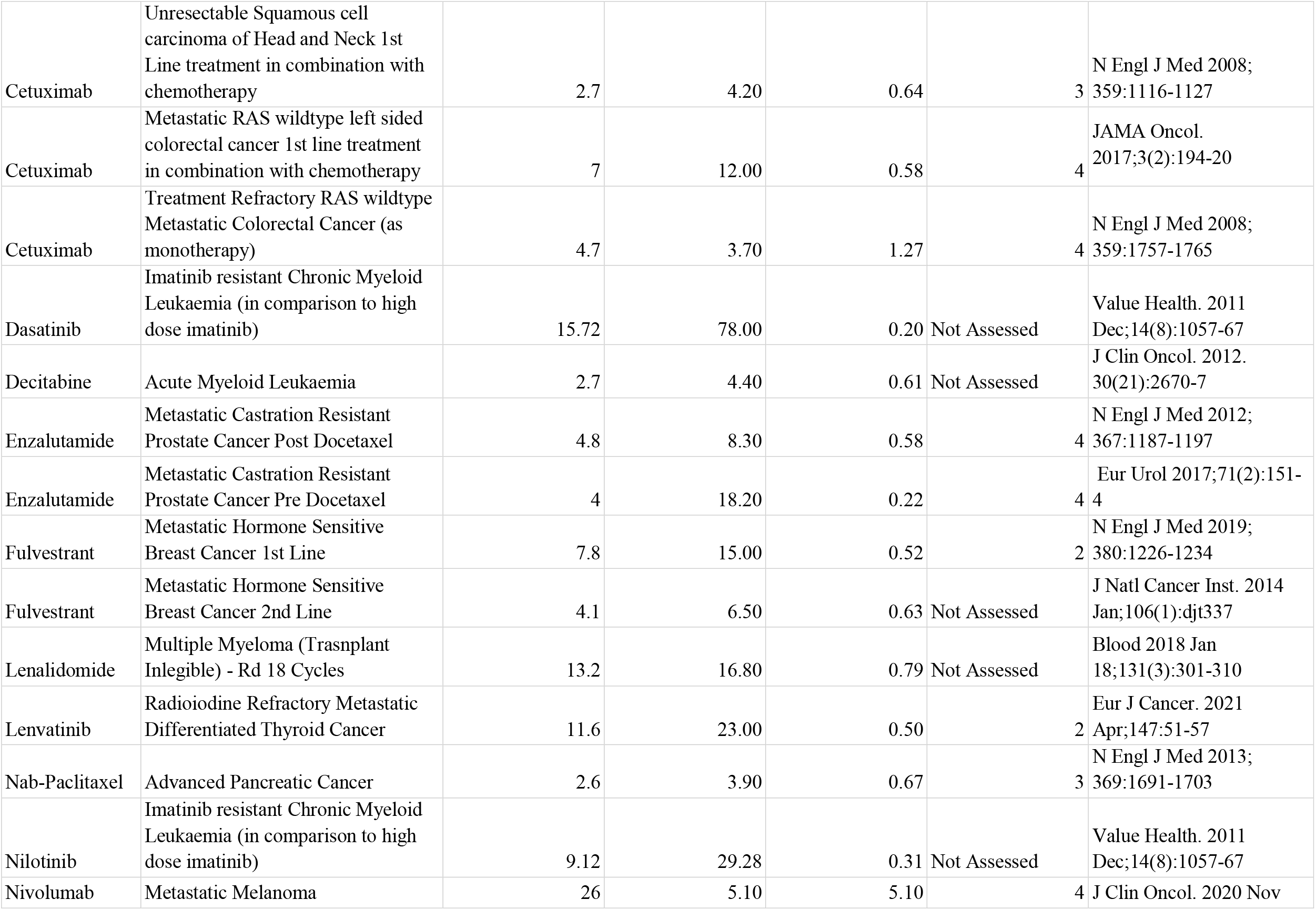
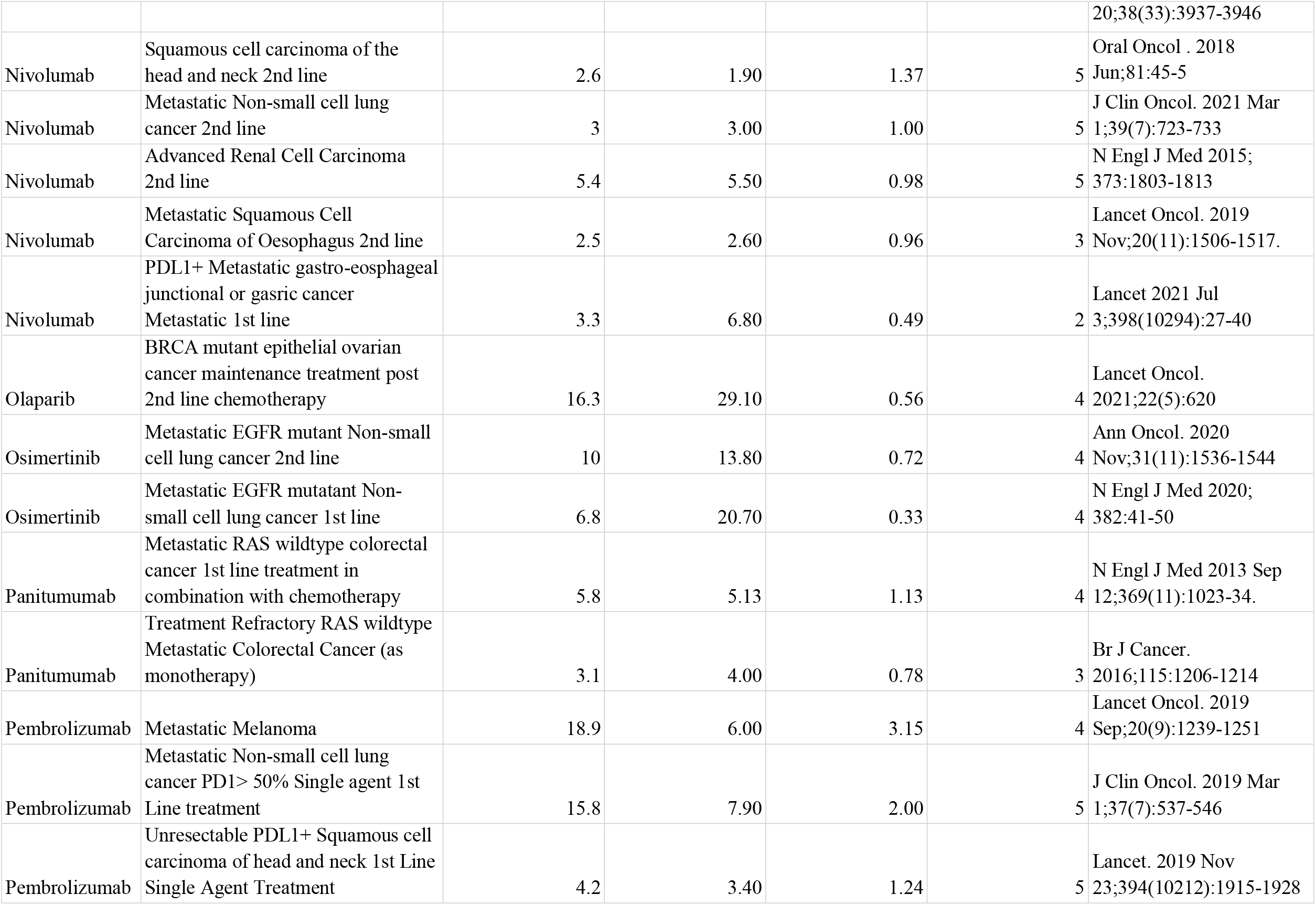
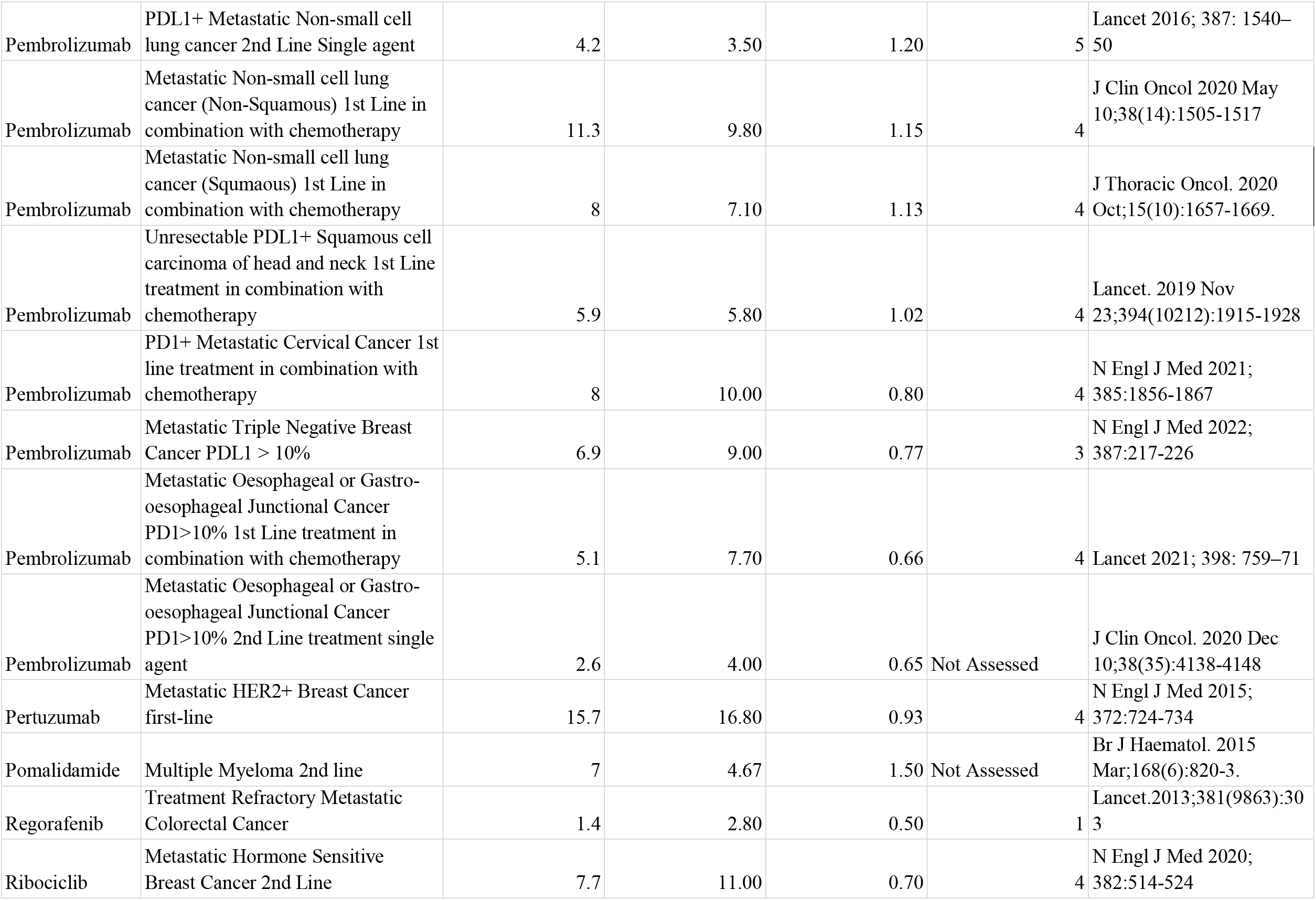
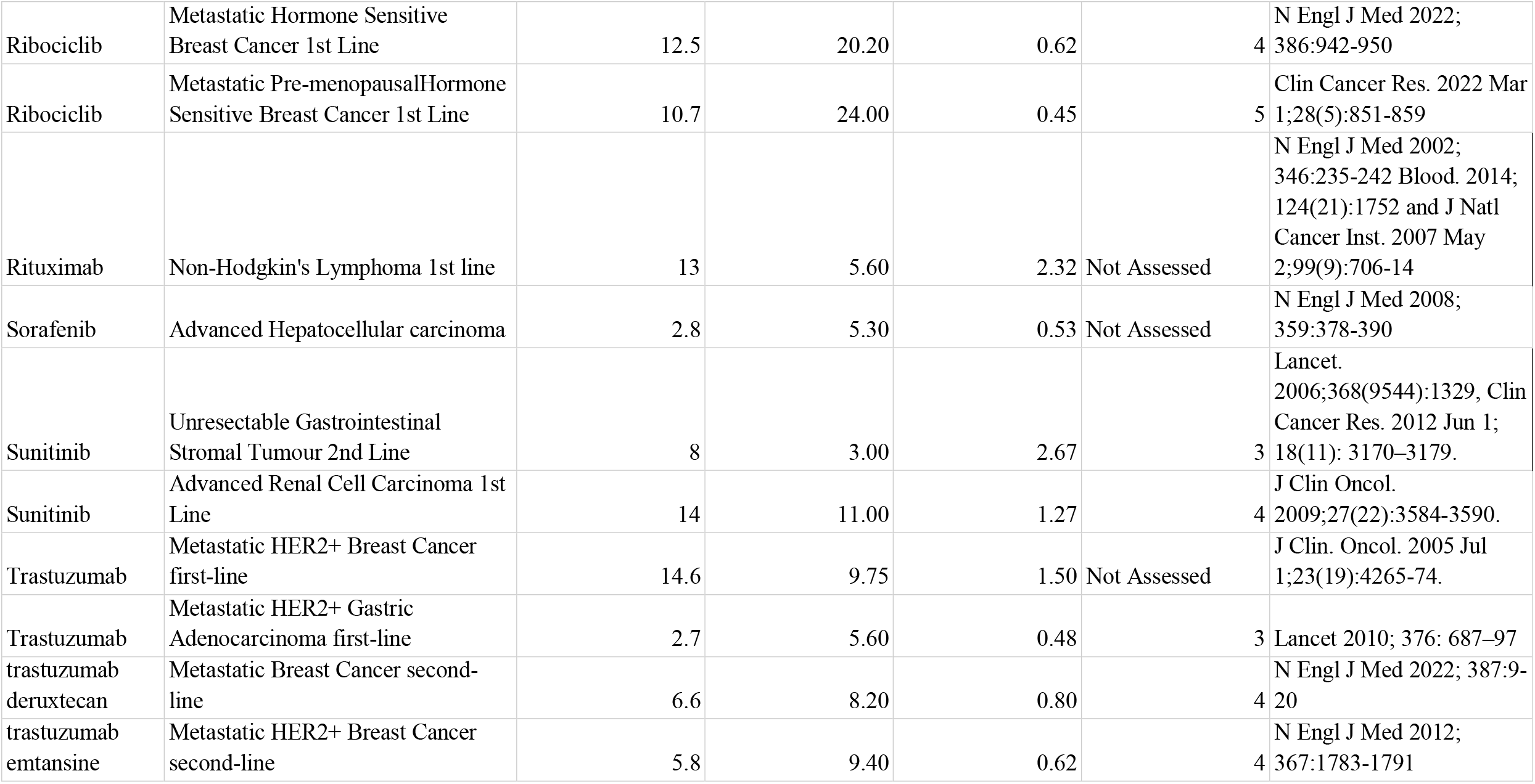
Overall survival gain per unit treatment time (Full Dataset)

**Supplementary Figure S1.**
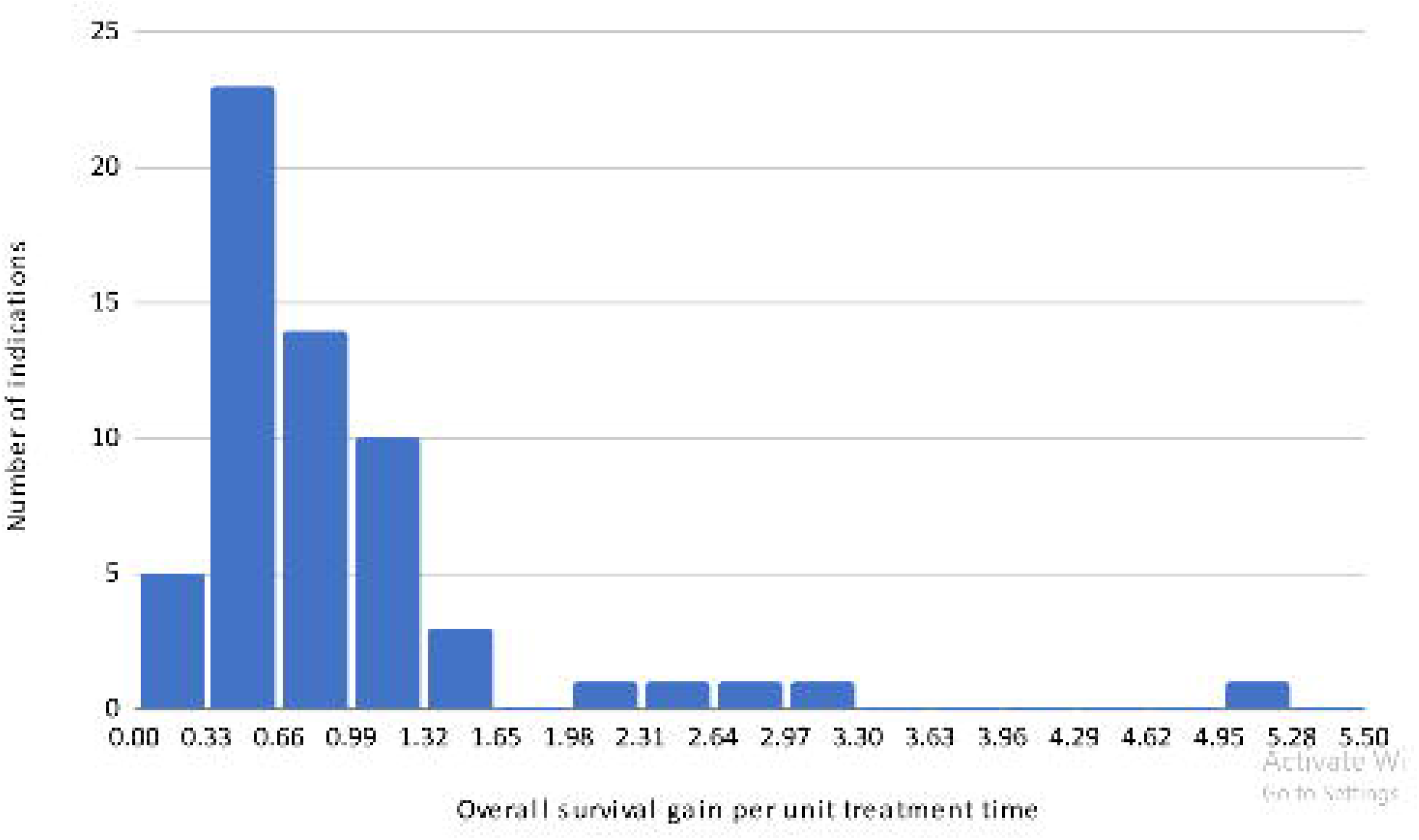
Distribution of indications against survival gain per unit treatment time.

